# Tattooing and cervical cancer risk: an unexpected finding from a negative control analysis in the CRABAT cohort

**DOI:** 10.1101/2025.09.24.25336531

**Authors:** Rachel D McCarty, Liacine Bouaoun, Marie Zins, Marcel Goldberg, Céline Ribet, Sofiane Kab, Tarik Gheit, Khaled Ezzedine, Joachim Schüz, Milena Foerster

**Affiliations:** Environment and Lifestyle Epidemiology Branch, International Agency for Research on Cancer, World Health Organization, Lyon, France; Université Paris Cité, Université Paris-Saclay, UVSQ, INSERM, Epidemiological Population Cohorts Unit (UMS 011), France; Université Paris Cité and Université Sorbonne Paris Nord, Inserm, INRAE, Centre for Research in Epidemiology and Statistics (CRESS), Paris, France; Centre d’Epidémiologie Clinique, Hôpital Hôtel Dieu, AP-HP, Paris, France; Early Detection, Prevention, and Infections Branch, International Agency for Research on Cancer, World Health Organization, Lyon, France; Department of Dermatology, Henri-Mondor University Hospital, Créteil, France; Epidemiology in Dermatology and Evaluation of Therapeutics (EpiDermE), EA 7379, Paris-Est Créteil University, Créteil, France

**Author notes:** **Corresponding author:** Rachel D McCarty, International Agency for Research on Cancer, 25 avenue Tony Garnier, 69007 Lyon, France.

**Keywords:** tattooing, cervical cancer, unmeasured confounding, confounding assessment, latent class analysis, negative control

## Abstract

**Background:** Tattooing is increasingly being investigated as a cancer risk factor, but is also strongly associated with smoking and other lifestyle factors that could confound such associations. To assess this potential confounding, we selected cervical cancer as a negative control, given that nearly all cases are attributable to HPV infection.

**Methods:** We analyzed data from 50,769 women in the Cancer Risk Associated with the Body Art of Tattooing (CRABAT) study nested within the French Constances cohort. We applied latent class analysis (LCA) to identify latent lifestyle profiles from sociodemographic, lifestyle, and health-related variables. Logistic regression models estimated associations between tattoos and cervical cancer, adjusting for confounders and latent lifestyle profile. Additional models were stratified by latent lifestyle profile.

**Results:** In minimally adjusted models, women with tattoos had a higher cervical cancer risk (odds ratio [OR]=1.54 [95% confidence interval [CI] 1.15–2.05]). Associations were stronger for larger tattooed surface area and longer time since tattooing. Associations were attenuated but persisted after adjusting for known confounders (1.33 [0.99–1.77]), and after additionally adjusting for latent lifestyle profile (1.29 [0.95–1.73]). Stratified models showed strong variations, with ORs ranging from 0.99 [0.37–2.61] in the younger profile to 2.56 [1.23–5.34] in the socially-advantaged profile.

**Conclusions:** Despite being selected as a negative control, elevated cervical cancer risk among tattooed women persisted even after comprehensive confounding adjustment. Stronger associations were observed with heavier tattoo exposure. This raises the possibility of a true association involving tattoo-related immune effects on HPV clearance as a potential biological mechanism.

## Background

Tattooing is being studied as a possible cancer risk factor due to lifelong exposure to tattoo ink components, some of which are classified as carcinogens.^1^ Initial studies have reported positive associations between tattooing and risk of hematologic cancers^2,3^ and mixed findings for skin cancers.^4–8^ While these studies adjusted for some risk factors like smoking, tattooing is highly correlated with many other sociodemographic and lifestyle factors that are difficult to comprehensively adjust for.^9,10^ First analyses of our Cancer Risk Associated with the Body Art of Tattooing (CRABAT) cohort showed clear differences between the tattooed and non-tattooed population for many established cancer risk factors such as education, income, and smoking, in addition to hepatitis B (HBV)/hepatitis C (HCV) infections and related sexual risk factors.^11^ These relationships may further be interdependent and time-changing, and multivariable adjustment may be insufficient to fully account for complex lifestyle-related confounding in the tattoo-cancer relationship, leaving residual confounding that may bias results.

We examined confounding affecting tattooing and cancer risk in the CRABAT study, using cervical cancer as an example. Cervical cancer was selected as a negative control because its etiology is well established; nearly all cases are attributable to cervical infection with mucosal human papillomavirus (HPV).^12^ Given this near-universal HPV etiology, we hypothesized that any observed associations between tattooing and cervical cancer would largely be explained by lifestyle confounding.

To explore how tattoo-cervical cancer associations might vary across distinct lifestyle patterns, we applied latent class analysis (LCA) to group women into profiles based on patterns of sociodemographic, lifestyle, and health-related variables. LCA uses probabilistic modeling to identify unobserved subgroups, or latent classes, within a population based on patterns of observed variables.^13^ This approach can be useful when distinct, unmeasured patterns among correlated variables are suspected and has been previously applied to help disentangle complex confounding, e.g., in studies of mobile phone use and radiation exposure.^14^ Rather than adjusting for a large number of individual, correlated variables, which can lead to sparse data, collinearity, and overfitting, LCA summarizes these variables into a smaller number of interpretable lifestyle profiles, allowing us to examine whether tattoo-cervical cancer associations were consistent across women with differing lifestyle patterns.

## Methods

### Study population

The French Constances cohort includes almost 220,000 participants, sampled to reflect the sociodemographic characteristics of the metropolitan French population.^15^ The cohort collects extensive data on occupational and social factors as well as health outcomes, and was established as a research infrastructure for the epidemiologic research community.^16^ Individuals were randomly sampled from adults aged 18 to 69, insured through the French General Health Insurance Fund, which covers approximately 85% of the population.^17^ Baseline data were collected via self-administered questionnaires between 2012 and 2020.^17^ In addition to follow-up questionnaires completed each year, Constances receives medical data through linkage to the French national health database (Système National des Données de Santé, SNDS) which contains records of all medical services reimbursed by the national public health insurance system.^11^ Written informed consent was obtained from all participants prior to their inclusion in the study.

CRABAT consists of over 110,000 Constances participants. As previously described,^11^ during the 2020–2021 follow-up, participants were asked “Do you have a tattoo?” Respondents to this question were enrolled in CRABAT. Between July and December 2023, individuals with tattoos were sent the validated Epidemiological Tattoo Assessment Tool (EpiTAT) questionnaire, which collected more detailed information on tattoo characteristics.^18^ We utilized a retrospective design as tattoo exposure was ascertained via self-report in 2020–2023, while cervical cancer diagnoses spanned 2005–2021, meaning diagnosis preceded exposure ascertainment for most cases.

### Tattoo exposure data

Data on ever receiving a tattoo (yes/no) were obtained from the 2020–2021 questionnaire. Other tattoo variables retrieved from the 2023 EpiTAT questionnaire were: total tattooed body surface area, timing of tattoos, and the context in which tattoos were received (i.e., by an experienced artist in a tattoo studio; by an experienced artist in other circumstances; or by a non-experienced person). For statistical analyses categories were collapsed to tattoos received by a professional artist in a studio versus other. Because participants could have multiple tattoos acquired at different times, tattoo timing was assessed using multiple-choice categories: within the last year, 1–5 years ago, 5–10 years ago, 10–15 years ago, more than 15 years ago. The midpoint of the earliest indicated range was used to estimate date of first tattoo. For those reporting more than 15 years ago, date of first tattoo was set to 20 years prior to questionnaire completion. Women who received their first tattoo the year of or after cancer diagnosis were considered non-exposed to ensure that exposure preceded the outcome (n=8). Women with tattoos missing detailed timing data (n=31, 41%) were retained in the ever/never tattoo variable but could not be evaluated for tattoo timing relative to diagnosis.

### Outcome data

The study population was restricted to women below age 70 because of the very low tattoo prevalence in older age groups. The outcome cervical cancer diagnosis was ascertained through linkage to the SNDS database. Eligible diagnoses were identified using ICD-10 codes, specifically C53 (malignant neoplasm of the cervix) and D06 (carcinoma in situ of the cervix). We excluded women without SNDS data (n=2,809) and women diagnosed with cervical cancer prior to 2005 as this was more than 15 years prior to the first tattoo questions (n=11) (Figure 1). A total of 50,769 women were included in analyses, 7,881 (16%) with tattoos and 42,888 (84%) without tattoos.

**Figure 1.**
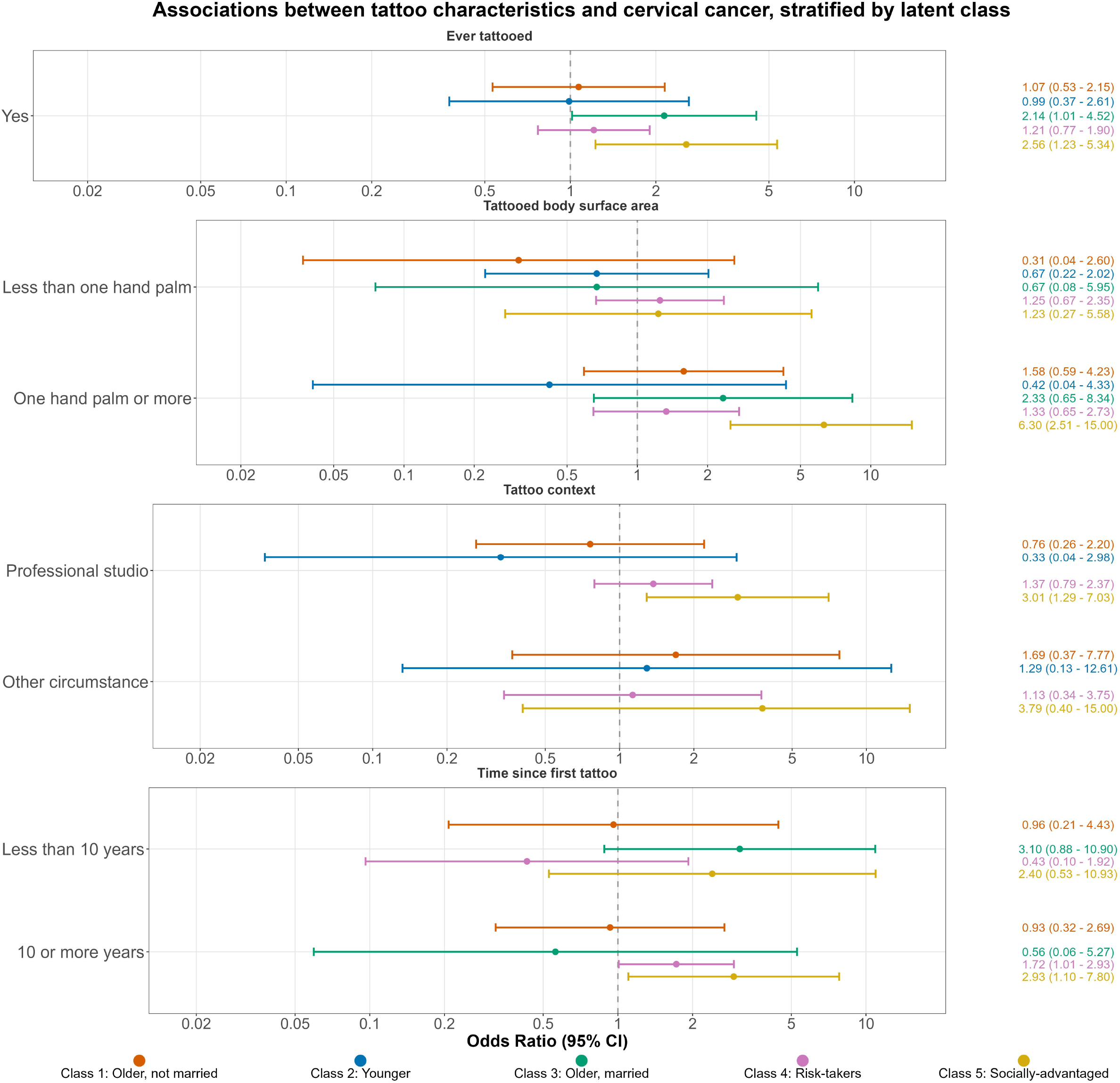
Flowchart of participant inclusion and exclusion

Cases were defined as those diagnosed with cervical cancer between 2005 and 2021. Women who were diagnosed with cervical cancer after 2021 (n=81) were classified as non-cases, as their diagnosis fell outside of the study period.

### Sociodemographic, lifestyle and health-related covariates

Relevant covariates were selected *a priori* using a directed acyclic graph (DAG) to identify variables plausibly associated with both tattooing and cervical cancer risk which was created using the DAGitty browser-based tool (dagitty.net) (Supplemental Figure 1). Based on prior literature, we adjusted for variables such as healthcare access, HPV vaccination uptake, and number of sexual partners, as each has been shown to differ between tattooed and non-tattooed individuals and is associated with cervical cancer risk.^9,11^

Sociodemographic, lifestyle, and health-related covariates used for latent profile model fitting and logistic regression were primarily obtained from the Constances baseline and 2020/2021 questionnaires. In general, the variable assessed closest to the 2020/2021 follow-up was used. However, if a more detailed assessment was available in an earlier questionnaire (e.g., the alcohol use score in the baseline questionnaire), that version was used instead of less detailed follow-up data. HPV vaccination and cervical smears were obtained from SNDS data between 2007 (the first-year data are available) and 2021.

Details on the questionnaire-derived covariates, including the specific questionnaire and answer categories, are provided in Supplemental Table 1.

### Statistical analysis

Demographic characteristics of tattooed and non-tattooed women were described using frequency distributions and standardized mean differences (SMDs), a common measure of effect size in observational studies. We considered an SMD of 0.20 or greater as indicative of a meaningful difference. The statistical modeling analysis was comprised of two parts. First, latent lifestyle profiles of women sharing similar sociodemographic, lifestyle, and health-related patterns were identified using LCA analysis. Second, we examined how associations between tattoos and cervical cancer varied when accounting for latent lifestyle profile.

### LCA

Initially all sociodemographic, lifestyle, and health-related covariates were considered in the LCA models. However, after examining the distribution of the variables across profiles, the final variables included were tobacco smoking, education, cannabis use, physical activity, number of lifetime sexual partners, age, body mass index (BMI), alcohol use, monthly income, marital status, general health score, depression, and rural/urban residence. The variables that were not included (healthcare access, HPV vaccination, and regular Pap smears) were retained in the logistic regression models as they were key confounders in our DAG.

We implemented LCA models with 2–8 latent lifestyle profiles and determined the optimal number of profiles based on model goodness-of-fit criteria, using the Akaike Information Criterion (AIC) and the Bayesian Information Criterion (BIC) while also considering interpretability. Item response probabilities were calculated for each response variable category within each latent lifestyle profile. To improve readability of results, we assigned descriptive labels to each latent lifestyle profile based on the predominant characteristics of women within that profile. These labels are intended as general summaries rather than precise definitions. Individual posterior probabilities of profile membership were calculated for each participant, and participants were assigned to the profile for which they had the highest posterior membership probability.

### Logistic regression models

Logistic regression was used to estimate odds ratios (ORs) and 95% confidence intervals (CIs) for associations between tattoo exposures and cervical cancer risk. We fit three models: A minimally adjusted model (Model 1) adjusted for age, ever smoking, and education; (Model 2) further adjusted for number of lifetime sexual partners, HPV vaccination, healthcare access, and regular Pap smears; and (Model 3) further adjusted for latent profile using the largest profile as the reference group. Covariates used in LCA were retained in regression models to address residual confounding within profiles. We also fit models stratified by latent profile, adjusted for age, number of lifetime sexual partners, and regular Pap smears. We calculated E-values^19,20^ to assess the potential impact of residual and unmeasured confounding on our fully adjusted models.

### Sensitivity analyses

We examined association between profile membership and cervical cancer risk excluding and including tattooed women; and between profile membership and ever receiving a tattoo using logistic regression. Models were also fit restricted to women receiving regular Pap smears.

To explore associations in a retrospective cohort approach, we also conducted Cox proportional hazards regression with age as the time scale and left truncation at enrollment. Tattoo exposure was modeled as ever/never (Approach 1) and as a time-varying covariate (Approach 2).

All analyses were performed in R (v4.3.1; R core team; Vienna, Austria) and a two-sided p-value < 0.05 was considered statistically significant.

## Results

### Demographics of tattooed and non-tattooed women

Among 50,769 women included in the analyses, 7,881 (16%) reported having a tattoo and 42,888 (84%) reported no tattoos. Tattooed women were younger, had lower education and income, were less likely to be married, and more likely to have smoked, used cannabis, and report more lifetime sexual partners (Table 1). Tattooed women were also more likely to have received at least one HPV vaccine dose, largely due to their younger age. Groups were well balanced in terms of alcohol use, BMI, and Pap smear uptake.

**Table 1.** Demographic characteristics of women under age 70 within the CRABAT cohort, overall and by tattoo status.

|  | Total<br>(n=50,769) |  | Tattooed<br>(n=7,881) |  | Not tattooed<br>(n=42,888) |  | Standardized<br>Mean<br>Difference (SMD) |
| --- | --- | --- | --- | --- | --- | --- | --- |
|  | n | (%) | n | (%) | n | (%) |  |
| **Age\*** |  |  |  |  |  |  | 0.70 |
| <30 | 1406 | (3%) | 482 | (6%) | 924 | (2%) |  |
| 30-39 | 8660 | (17%) | 2412 | (31%) | 6248 | (15%) |  |
| 40-49 | 13070 | (26%) | 2571 | (33%) | 10499 | (24%) |  |
| 50-59 | 13172 | (26%) | 1665 | (21%) | 11507 | (27%) |  |
| 60-69 | 14461 | (28%) | 751 | (10%) | 13710 | (32%) |  |
| **Education** |  |  |  |  |  |  | 0.30 |
| No diploma | 7500 | (15%) | 1392 | (18%) | 6108 | (14%) |  |
| High school diploma | 7684 | (15%) | 1734 | (22%) | 5950 | (14%) |  |
| College, Bachelor's diploma | 21539 | (42%) | 3235 | (41%) | 18304 | (43%) |  |
| Masters diploma or higher | 13228 | (26%) | 1406 | (18%) | 11822 | (28%) |  |
| Missing | 818 | (2%) | 114 | (1%) | 704 | (2%) |  |
| **Monthly income, in euros** |  |  |  |  |  |  | 0.38 |
| <1500 | 4104 | (8%) | 1077 | (14%) | 3027 | (7%) |  |
| >=1500-<2800 | 12200 | (24%) | 2270 | (29%) | 9930 | (23%) |  |
| >=2800-<4200 | 15890 | (31%) | 2588 | (33%) | 13302 | (31%) |  |
| >=4200 | 15071 | (30%) | 1368 | (17%) | 13703 | (32%) |  |
| No answer | 3504 | (7%) | 578 | (7%) | 2926 | (7%) |  |
| **Marital status** |  |  |  |  |  |  | 0.27 |
| Unmarried | 12820 | (25%) | 2749 | (35%) | 10071 | (23%) |  |
| Married/civil partnership | 31494 | (62%) | 4080 | (52%) | 27414 | (64%) |  |
| Was married | 6430 | (13%) | 1045 | (13%) | 5385 | (13%) |  |
| Missing | 25 | (0%) | 7 | (0%) | 18 | (0%) |  |
| **Ever smoking** |  |  |  |  |  |  | 0.36 |
| Yes | 23085 | (45%) | 4724 | (60%) | 18361 | (43%) |  |
| No | 26193 | (52%) | 2933 | (37%) | 23260 | (54%) |  |
| Missing | 1491 | (3%) | 224 | (3%) | 1267 | (3%) |  |
| **Cannabis use** |  |  |  |  |  |  | 0.52 |
| Never | 31515 | (62%) | 3397 | (43%) | 28118 | (66%) |  |
| Former | 15265 | (30%) | 3583 | (45%) | 11682 | (27%) |  |
| Current | 1893 | (4%) | 676 | (9%) | 1217 | (3%) |  |
| Missing | 2096 | (4%) | 225 | (3%) | 1871 | (4%) |  |
| **Alcohol use** |  |  |  |  |  |  | 0.12 |
| Abstainer | 11193 | (22%) | 1633 | (21%) | 9560 | (22%) |  |
| Abuse or dependence | 5181 | (10%) | 1058 | (13%) | 4123 | (10%) |  |
| Moderate use | 33479 | (66%) | 5032 | (64%) | 28447 | (66%) |  |
| Missing | 916 | (2%) | 158 | (2%) | 758 | (2%) |  |
| **BMI** |  |  |  |  |  |  | 0.06 |
| Underweight | 2215 | (4%) | 361 | (5%) | 1854 | (4%) |  |
| Normal weight | 31326 | (62%) | 4722 | (60%) | 26604 | (62%) |  |
| Overweight | 11380 | (22%) | 1793 | (23%) | 9587 | (22%) |  |
| Obese | 5353 | (11%) | 930 | (12%) | 4423 | (10%) |  |
| Missing | 495 | (1%) | 75 | (1%) | 420 | (1%) |  |
| **Number of lifetime sexual partners** |  |  |  |  |  |  | 0.33 |
| None | 16285 | (32%) | 2404 | (31%) | 13881 | (32%) |  |
| One | 794 | (2%) | 89 | (1%) | 705 | (2%) |  |
| Two to five | 8434 | (17%) | 773 | (10%) | 7661 | (18%) |  |
| Six or more | 12263 | (24%) | 2722 | (35%) | 9541 | (22%) |  |
| No answer | 12993 | (26%) | 1893 | (24%) | 11100 | (26%) |  |
| **Regular Pap smears\*\*** |  |  |  |  |  |  | 0.05 |
| Yes | 42619 | (84%) | 6732 | (85%) | 35887 | (84%) |  |
| No | 8150 | (16%) | 1149 | (15%) | 7001 | (16%) |  |
| **At least one dose of HPV vaccine** |  |  |  |  |  |  | 0.21 |
| Yes | 3014 | (6%) | 837 | (11%) | 2177 | (5%) |  |
| No | 47755 | (94%) | 7044 | (89%) | 40711 | (95%) |  |

\*Age in 2022 (the last year of cancer diagnoses)
\*\*At least 2 pap smears between 2008-2021
Abbreviations: Body mass index (BMI), human papillomavirus (HPV)
Note: We considered an SMD of 0.20 or greater as indicative of a meaningful difference

### Description of latent profiles

We selected a five-profile model as it provided an optimal balance between model goodness-of-fit, well-defined lifestyle pattern groups, and maintaining sufficient sample sizes for stratified analyses.

Item response probabilities for the sociodemographic, lifestyle, and health-related variables within each latent profile are shown in Table 2. The five latent profiles were characterized as follows: Profile 1 (“Older, not married”) included older, unmarried or previously married women who were unlikely to have used cannabis; Profile 2 (“Younger”) included women primarily under age 50, unlikely to have smoked tobacco or married; Profile 3 (“Older, married”) comprised older, married women unlikely to use cannabis; these women also tended to have lower educational attainment and lived in more rural areas; Profile 4 (“Risk-takers”) included women who were the most likely to smoke tobacco, use cannabis, meet criteria for alcohol abuse or dependence, and report a higher number of sexual partners; Profile 5 (“Socially-advantaged”) included women with higher income and education levels, and a lower likelihood of smoking tobacco or using cannabis. Profile membership was distributed as: 14% in Profile 1, 12% in Profile 2, 20% in Profile 3, 22% in Profile 4, and 31% in Profile 5, with Profile 5 as the reference group. Demographic and lifestyle characteristics are summarized across latent profiles in Supplemental Table 2.

**Table 2.** Item response probabilities for sociodemographic, lifestyle, and health-related variables within each latent lifestyle profile.

|  |  | Latent lifestyle profile |  |  |  |  |
| --- | --- | --- | --- | --- | --- | --- |
|  |  | Profile 1<br>Older, not<br>married | Profile 2<br>Younger | Profile 3<br>Older,<br>married | Profile 4<br>Risk-takers | Profile 5<br>Socially-advantaged |
| **Class membership probability** |  | 0.14 | 0.12 | 0.20 | 0.22 | 0.31 |
| **Item Response Probabilities** |  |  |  |  |  |  |
| Variable | Category |  |  |  |  |  |
| Smoking | Never | 0.48 | 0.72 | 0.55 | 0.08 | 0.78 |
|  | Former | 0.38 | 0.16 | 0.38 | 0.69 | 0.20 |
|  | Current | 0.15 | 0.13 | 0.06 | 0.23 | 0.02 |
| Education | No diploma | 0.27 | 0.05 | 0.38 | 0.05 | 0.00 |
|  | High school diploma | 0.20 | 0.27 | 0.28 | 0.10 | 0.03 |
|  | Bachelor's diploma | 0.42 | 0.38 | 0.33 | 0.48 | 0.52 |
|  | Master's diploma or higher | 0.10 | 0.30 | 0.01 | 0.37 | 0.45 |
| Cannabis use | Never | 0.80 | 0.54 | 0.95 | 0.05 | 0.84 |
|  | Former | 0.19 | 0.35 | 0.05 | 0.85 | 0.15 |
|  | Current | 0.02 | 0.11 | 0.00 | 0.10 | 0.00 |
| Physical activity | Low | 0.09 | 0.12 | 0.07 | 0.08 | 0.08 |
|  | Moderate | 0.63 | 0.71 | 0.60 | 0.69 | 0.68 |
|  | High | 0.28 | 0.17 | 0.33 | 0.23 | 0.25 |
| Lifetime sexual partners | None | 0.27 | 0.33 | 0.29 | 0.26 | 0.42 |
|  | 1 | 0.02 | 0.10 | 0.00 | 0.00 | 0.00 |
|  | 2-5 | 0.04 | 0.14 | 0.30 | 0.03 | 0.25 |
|  | 6 or more | 0.29 | 0.28 | 0.06 | 0.53 | 0.15 |
|  | No answer | 0.38 | 0.14 | 0.34 | 0.18 | 0.18 |
| Age | <30 | 0.00 | 0.23 | 0.00 | 0.00 | 0.00 |
|  | 30-39 | 0.02 | 0.57 | 0.03 | 0.23 | 0.16 |
|  | 40-49 | 0.15 | 0.17 | 0.12 | 0.40 | 0.36 |
|  | 50-59 | 0.36 | 0.03 | 0.31 | 0.24 | 0.29 |
|  | 60-69 | 0.48 | 0.00 | 0.55 | 0.14 | 0.19 |
| BMI | Underweight | 0.03 | 0.08 | 0.02 | 0.05 | 0.05 |
|  | Normal weight | 0.54 | 0.66 | 0.50 | 0.68 | 0.71 |
|  | Overweight | 0.27 | 0.17 | 0.31 | 0.20 | 0.18 |
|  | Obese | 0.16 | 0.09 | 0.17 | 0.07 | 0.05 |
| Alcohol use | Abstainer | 0.29 | 0.30 | 0.25 | 0.11 | 0.22 |
|  | Abuse or dependence | 0.08 | 0.13 | 0.06 | 0.22 | 0.05 |
|  | Moderate use | 0.63 | 0.58 | 0.68 | 0.67 | 0.73 |
| Monthly income in euros | <1500 | 0.24 | 0.25 | 0.02 | 0.04 | 0.00 |
|  | ≥1500-<2800 | 0.55 | 0.38 | 0.25 | 0.21 | 0.06 |
|  | ≥2800-<4200 | 0.15 | 0.18 | 0.45 | 0.37 | 0.32 |
|  | ≥4200 | 0.02 | 0.06 | 0.19 | 0.36 | 0.58 |
|  | No answer | 0.04 | 0.13 | 0.10 | 0.03 | 0.04 |
| Marital / relationship status | Never married | 0.42 | 0.88 | 0.01 | 0.27 | 0.09 |
|  | Married/civil partnership | 0.01 | 0.11 | 0.96 | 0.63 | 0.85 |
|  | Separated, divorced, or widow(er) | 0.58 | 0.00 | 0.03 | 0.09 | 0.06 |
| General health (score) | Good (1-2) | 0.43 | 0.58 | 0.46 | 0.56 | 0.67 |
|  | Moderate (3-4) | 0.44 | 0.34 | 0.44 | 0.36 | 0.29 |
|  | Poor (≥5) | 0.13 | 0.07 | 0.10 | 0.08 | 0.04 |
| Depression | Yes | 0.33 | 0.09 | 0.22 | 0.19 | 0.11 |
|  | No | 0.67 | 0.91 | 0.78 | 0.81 | 0.89 |
| Residence | Suburban | 0.33 | 0.31 | 0.36 | 0.30 | 0.38 |
|  | Central city | 0.44 | 0.44 | 0.21 | 0.45 | 0.38 |
|  | Isolated city | 0.07 | 0.06 | 0.11 | 0.06 | 0.07 |
|  | Rural | 0.16 | 0.18 | 0.32 | 0.19 | 0.18 |

Note: All percentages are rounded to the nearest whole number.
Dark gray cells indicate high (0.70–1.00), light gray cells indicate medium (0.30–0.69), and white cells indicate low (0.00–0.29).
Abbreviation: Body mass index (BMI)

### Tattoo exposures and risk of cervical cancer

Women with tattoos had a 54% higher risk of cervical cancer (OR=1.54 [95% CI 1.15– 2.05]) (Table 3, Model 1). This association was attenuated after additional adjustment for number of lifetime sexual partners, HPV vaccination, healthcare access, and regular Pap smears (OR=1.33 [95% CI 0.99–1.77]) (Model 2) and after further adjustment for latent lifestyle profile (OR=1.29 [95% CI 0.95–1.73]) (Model 3), though point estimates remained elevated in both models.

**Table 3.** Associations between tattoos and cervical cancer risk.

|  | **Cancer cases**<br>(n=320) |  | **Non-cases**<br>(n=50,449) |  | Model 1 | Model 2 | Model 3 |
| --- | --- | --- | --- | --- | --- | --- | --- |
|  | n | % | n | % | OR (95% CI) | OR (95% CI) | OR (95% CI) |
| **Ever tattooed** |  |  |  |  |  |  |  |
| No | 244 | 76% | 42,644 | 85% | Ref | Ref | Ref |
| Yes | 76 | 24% | 7,805 | 15% | 1.54 (1.15–2.05) | 1.33 (0.99–1.77) | 1.29 (0.95–1.73) |
| **Tattooed body surface area** |  |  |  |  |  |  |  |
| Never tattooed | 244 | 76% | 42,644 | 85% | Ref | Ref | Ref |
| < One hand palm | 21 | 7% | 2,523 | 5% | 1.33 (0.81–2.07) | 1.16 (0.71–1.80) | 1.03 (0.60–1.65) |
| One hand palm or more | 26 | 8% | 2,244 | 4% | 1.93 (1.23–2.91) | 1.63 (1.03–2.46) | 1.58 (0.99–2.43) |
| **Tattoo context** |  |  |  |  |  |  |  |
| Never tattooed | 244 | 76% | 42,644 | 85% | Ref | Ref | Ref |
| Only tattooed by professional artist inside studio | 38 | 12% | 3,931 | 8% | 1.57 (1.08–2.24) | 1.37 (0.94–1.94) | 1.28 (0.85–1.61) |
| Tattooed in other circumstance | 8 | 3% | 782 | 2% | 1.63 (0.73–3.13) | 1.33 (0.60–2.57) | 1.24 (0.52–2.49) |
| **Time since first tattoo** |  |  |  |  |  |  |  |
| Never tattooed | 244 | 76% | 42,644 | 85% | Ref | Ref | Ref |
| Less than 10 years | 10 | 3% | 1,772 | 4% | 0.96 (0.50–1.74) | 0.89 (0.44–1.62) | 0.86 (0.40–1.60) |
| 10 or more years | 35 | 11% | 2,930 | 6% | 1.89 (1.28–2.71) | 1.56 (1.06–2.25) | 1.43 (0.94–2.11) |
| Missing data on timing of tattoo | 31 | 10% | 3,103 | 6% | - | - | - |

Model 1 adjusted for age, ever smoking, and education
Model 2 adjusted for age, ever smoking, education, number of lifetime sexual partners, HPV vaccination, healthcare access, and regular Pap smears
Model 3 adjusted for age, ever smoking, education, latent lifestyle profile, number of lifetime sexual partners, HPV vaccination, healthcare access, and regular Pap smears
Variable categories: age (< 30, 30–39, 40–49, 50–59, 60–69); ever (tobacco) smoking (never, former, current); education (no diploma, high school diploma, bachelor's diploma, master's diploma or higher); number of lifetime sexual partners (none, one, two to five, six or more, no answer); HPV vaccination (yes/no); healthcare access (at least one instance of forgoing healthcare due to financial problems in the past year or not); regular Pap smears (received two or more Pap smears during the 2008–2021 period or not).

The highest risk was observed among women with a tattooed body surface area of one hand palm or more and those tattooed 10 or more years prior to questionnaire completion, each associated with an approximately 90% increased risk compared with never-tattooed women (Model 1). Both associations attenuated to approximately 60% after adjustment for additional confounders (Model 2), and after further adjustment for latent lifestyle profile, to 58% for body surface area of one hand palm or more and 43% for tattoos received 10 or more years prior (Model 3).

### Tattoo exposures and risk of cervical cancer stratified by latent profile

Point estimates varied across latent lifestyle profiles, though most estimates were statistically imprecise (Figure 2). The strongest association with ever receiving a tattoo, with roughly a two and a half-fold increased risk, was observed within the socially-advantaged group (Profile 5, OR=2.56 [95% CI 1.23–5.34]) compared with never getting tattooed, followed by the older, married group (Profile 3, OR=2.14 [95% CI 1.01–4.52]). The association was weaker within the risk-taker group (Profile 4, OR=1.21 [95% CI 0.77–1.90]) and close to null within the older, not married (Profile 1, OR=1.07 [95% CI 0.53–2.15]) and the younger group (Profile 2, OR=0.99 [95% CI 0.37–2.61]).

**Figure 2.** Odds ratios and 95% confidence intervals for associations between tattoo characteristics and cervical cancer, by latent class

Having a tattooed body surface area of one hand palm or more was associated with more than a six-fold increased risk of cervical cancer in the socially-advantaged group (Profile 5, OR=6.30 [95% CI 2.51–15.00]) with positive but imprecise associations in the older, not married group (Profile 1, OR=1.58 [95% CI 0.59–4.23]) and the older, married group (Profile 3, OR=2.33 [95% CI 0.65–8.34]), and no clear pattern in the younger (Profile 2) and risk-taking groups (Profile 4).

Analyses of tattoo context (by a professional in a studio vs. other circumstance) were limited by small sample sizes, preventing clear patterns from emerging.

### Sensitivity analyses

Ever receiving a tattoo was associated with latent lifestyle profile with approximately 65% higher odds among the risk-taker group (Profile 4) and the older, not married group (Profile 1), compared with the socially-advantaged group as the referent group (Supplemental Table 3).

Compared to the socially-advantaged group, risk of cervical cancer was higher in all other latent lifestyle profiles, while risks were somewhat lower when including vs excluding tattooed individuals from the analysis in the younger and risk-taker profiles (Supplemental Table 4).

In analyses between tattoos and cervical cancer risk restricted to individuals receiving regular Pap smears, adjustment for HPV risk factors (Model 2) had little influence on the estimate while risks were slightly more attenuated when additionally adjusting for latent profiles (Model 3, Supplemental Table 5).

In time-to-event analyses, results were broadly consistent with primary logistic regression findings across both approaches and most latent profiles (Supplemental Tables 6 and 7). These findings support the robustness of the main results.

E-values for all associations within fully adjusted models ranged from 1.21–2.54 (Supplemental Table 8), indicating that a weak-to-moderate unmeasured confounder could explain away some findings. However complex interdependent confounding could collectively explain these associations at lower individual effect sizes than these thresholds suggest.

## Discussion

In this study, we selected cervical cancer as a negative control outcome to examine confounding in associations between tattooing and cancer, given that nearly all cervical cancer cases are attributable to HPV infection. Contrary to our expectations, we observed a persistent, elevated risk of cervical cancer among tattooed women that remained after adjustment for known confounders and latent lifestyle profiles. While attenuation across models suggests lifestyle confounding contributes to this association, the persistence of elevated risk, alongside stronger associations for larger tattoo surface area and longer tattoo duration, raises the possibility of a biological mechanism.

A growing body of evidence suggests that tattooing may have immunomodulatory effects beyond the tattooed skin; however, whether these effects have clinical relevance at distant mucosal sites remains unclear.^21,22^ To our knowledge, no direct association between tattoo exposure and cervical cancer has previously been established, and any proposed biological mechanism therefore remains speculative. One potential hypothesis involves the lymphatic transport and long-term accumulation of tattoo pigments in draining lymph nodes. Tattoo pigments are taken up predominantly by macrophages and can induce persistent inflammatory changes within draining lymph nodes; an experimental study in mice has also demonstrated macrophage toxicity and an acute systemic inflammatory response following tattooing.^21^ However, it remains unclear whether tattoo-associated immune modulation could extend beyond the lymphatic compartment and influence local immune responses in the genital tract, affecting clearance of persistent high-risk HPV infection and thereby contribute to cervical carcinogenesis. The same study in tattooed mice also found that tattooing differentially modulated vaccine-induced immune responses, reducing the antibody response to an mRNA-based SARS-CoV-2 vaccine while enhancing the response to a UV-inactivated influenza vaccine.^21^ Overall, whether tattoo-associated immune alterations influence HPV acquisition or persistence, HPV-specific immunity, responses to HPV vaccination, or progression from oncogenic HPV infection to cervical precancer and cancer remains unknown. Further mechanistic studies, together with well-designed epidemiological investigations, are therefore needed to determine whether the observed association reflects a causal relationship.

Stratified analyses revealed variation across latent profiles, providing insight into a confounding structure, supporting the role of residual confounding in the tattoo-cancer association. The strongest association was observed in the socially-advantaged profile, potentially reflecting greater correlation between tattooing and unmeasured HPV-related lifestyle factors in this group. In contrast, associations were weaker or near-null in the older not married, younger, and risk-taking profiles. One explanation could be that tattoo status does not distinguish individuals within these groups well, including on other cancer-related factors. Such patterns suggest that residual confounding varies meaningfully by lifestyle context, supporting our initial hypothesis that standard multivariable adjustment may be insufficient to fully capture this complexity.

### Strengths and limitations

Strengths of this study include the detailed exposure, sociodemographic, and lifestyle data which are available in CRABAT through its linkage to the French Constances cohort. The primary limitations of this study are the timing of tattoo exposure ascertainment in 2020– 2021 (with detailed tattoo characteristics collected in 2023), after most cervical cancers had already been diagnosed, and substantial missingness in detailed tattoo data, including timing of tattooing, which was absent for approximately 40% of the tattooed sample. This raises several concerns. First, if a substantial proportion of the women with missing tattoo data were tattooed after their diagnosis, reverse causation could contribute to the observed association. Second, women who died prior to this assessment, including from cervical cancer, were not represented in either exposure group, which may introduce survivorship bias. We were further limited by the lack of data on HPV infection status. Additional limitations include small sample sizes within stratified analyses, which affected the precision of estimates, and selection bias in our case definition, as 81 women diagnosed after the study period (20% of potential cases) were classified as non-cases, biasing estimates toward the null. There is also tension between overadjustment and residual confounding in our analytical approach. We addressed this by presenting models with different levels of adjustment. The Constances cohort has a participation rate of approximately 7.3%, with overrepresentation of more educated and higher socioeconomic status individuals relative to the general French population,^16^ which may limit the generalizability of our findings.

Furthermore, outcome ascertainment via administrative records may incompletely capture early-stage or screen-detected cervical cancers managed in outpatient settings. Despite these limitations, this study provides a novel application of LCA to examine how sociodemographic and lifestyle variables may affect associations between tattoos and cancer. Future work could incorporate methods that account for uncertainty in profile membership assignments (e.g., weighting by posterior probabilities) to improve robustness of the findings.

## Conclusions

We selected cervical cancer as a negative control for exploring associations between tattoos and cancer, anticipating that any observed association would be attributable to confounding. The unexpected persistence of elevated risk among tattooed women after comprehensive confounding adjustment, alongside stronger associations for larger tattoo surface area and longer tattoo duration, raises the possibility that biological factors beyond confounding may contribute to the observed association. Tattoo-induced immunomodulation is one potential biological hypothesis. However, the immune effects of tattooing at distant mucosal sites and on responses relevant to HPV acquisition and persistence remain unknown and largely unexplored. LCA revealed substantial heterogeneity in risk across lifestyle profiles, suggesting confounding varies meaningfully by lifestyle context. These findings highlight both the complexity of confounding in tattoo-cancer research and the need for dedicated studies of tattoo-related immune effects.

## Data availability

For data security and participant confidentiality, the data used in this research is hosted on the secured Centre d’Accès Sécurisé aux Données (CASD) platform. Access is strictly regulated. All data access requests must be formally addressed to the Constances and CRABAT study teams via the procedures outlined on the Constances website (constances.fr).

## Ethics approval

The Constances study was approved by the French Institute of Health institutional review board (IRB) (Opinion n°01-011, then n°21-842), and was authorized by the French Data Protection Authority (“Commission Nationale de l’Informatique et des Libertés”, CNIL) (Authorization #910486). The CRABAT study was approved by the International Agency for Research on Cancer (IARC) Ethic Committee (IEC 22-02) and was authorized by the Commission Nationale de l’Informatique et des Libertés (CNIL, #22015584).

## Disclaimer

Where authors are identified as personnel of the International Agency for Research on Cancer or the World Health Organization, the authors alone are responsible for the views expressed in this paper; the views do not necessarily represent the decisions, policy, or views of the International Agency for Research on Cancer or the World Health Organization.

## Acknowledgements

The authors thank the team of the Epidemiological Population Cohorts Unit in France (UMS 011 – Inserm, Universités Paris Cité, Paris Saclay, Versailles Saint-Quentin), which designed and manages the Constances cohort. They also acknowledge the French national health insurance fund (Caisse nationale d’assurance maladie, CNAM) and its Health examination centers for collecting a large part of the data, as well as the French national old-age insurance fund (Caisse nationale d’assurance vieillesse, CNAV) for their contribution to the establishment of the cohort, and ClinSearch for performing data quality control.

