## Supplementary material for "Tattooing and cervical cancer risk: an unexpected finding from a negative control analysis in the CRABAT cohort"

**Supplemental Table 1. Source and description of covariates from questionnaires used in latent class analysis and logistic regression models**

| **Variable** | **Question** | **Questionnaire answer categories** | **Answer categories used with different from questionnaire*** | **Questionnaire source** | **Role in analysis**** |
| --- | --- | --- | --- | --- | --- |
| **Sociodemographic variables** | | | | | |
| Education | What is the highest degree you have obtained? | 1 No diploma 2 Vocational training 3 High school diploma 4 College, bachelor's diploma 5 College, master's diploma or higher | No diploma/vocational training High school diploma College, bachelor's diploma College, master's diploma or higher | Baseline | LCA, adjustment |
| Income | What is the total amount of your household's net monthly income (i.e., the sum of the income of the people in your household or your own income if you live alone, regardless of the origin) | 1 Less than 450 € 2 450 € to less than 1000 € 3 1000 € to less than 1500 € 4 1500 € to less than 2100 € 5 2100 € to less than 2800 € 6 2800 € to less than 3200 € 7 4200 € or more 8 Don't know 9 Don't wish to answer | <1500 € ≥1500 to <2800 € ≥2800 to <4200 € ≥4200 € Don't know/don't wish to answer/missing | Baseline | LCA |
| Marital status | What is your family situation in terms of civil status? | 1 Single (never married) 2 Civil partnership 3 Married 4 Separated 5 Divorced 6 Widowed | Single (never married) Civil partnership/married Separated/divorced/widowed | 2020/2021 | LCA |
| Urban/rural residence | Geocode | Suburban Central city Isolated city Rural |  | Most recent residence based on each year of follow-up questionnaire starting in 2012 | LCA |
| Healthcare access | In the last 12 months, have you or your spouse had to forgo certain healthcare due to financial problems? | 1 Yes 2 No |  | 2020/2021 | LCA, adjustment |
| **Lifestyle variables** | | | | | |
| Tobacco smoking | During your life, have you ever used tobacco? | 1 Yes 2 No |  | 2020/2021 | LCA, adjustment |
| Cannabis | During your life, have you ever used cannabis (hashish, marijuana, grass, joint)? | 1 Yes 2 No |  | 2020/2021 | LCA |
| Physical activity (calculated from 3 questions) | Over the past 12 months, have you regularly traveled on foot, by bike, etc. (for work or not)? | 1 No (0 points) 2 Yes, less than 15 minutes each way (1 point) 3 Yes, 15 or more minutes each way (2 points) | Combined physical activity score was made from scoring each question.  The combined score was categorized as: Low = 0–1 Moderate = 2–4 High = 5 or more | 2020/2021 | LCA |
|  | Over the past 12 months have you regularly exercised (excluding commuting, DIY, gardening, and housework)? | 1 No (0 points) 2 Yes, less than 2 hours per week (1 point) 3 Yes, 2 or more hours per week (2 points) |  |  |  |
|  | During the last 12 months, have you regularly done DIY, gardening, or housework? | 1 No (0 points) 2 Yes, less than 2 hours per week (1 point) 3 Yes, 2 or more hours per week (2 points) |  |  |  |
| Sexual partners | During your life, with how many different partners have you had sexual intercourse? | Open-ended answer | NA/Never had sex 1 partner 2–5 partners 6 or more Prefer not to answer | Baseline | LCA, adjustment variable |
| Alcohol use (calculated from 3 questions) | How often do you usually consume alcoholic beverages? | 1 One to several times a week (if checked, without reponse to number of days = 3 points)  __ Indicate how many days (0.5–3 days = 3 points; 4 or more days = 4 points) 2 Two to three times per month (2 points) 3 One time per month or less (1 point) 4 Never (0 points) | Abstainers = Never consumed alcohol Neither abuse nor dependence = Combined score of ≤7 for men and ≤6 for women Abuse = Audit score >7 and ≤12 for men; and >6 and ≤11 for women Dependence = Audit score >12 for men and >11 for women | Original score was created at baseline; the score was updated with 2019 data when available | LCA |
|  | On days when you consume alcohol, how many standard alcoholic drinks do you drink on average during a day? | Open-ended answer (If number of drinks is: 0 to 2.5 = 0 points 2.5 to ≤4.5 = 1 point 4.5 to ≤6.5 = 2 points 6.5 to ≤8.5 = 3 points Greater than 8.5 = 4 points) |  |  |  |
|  | How often do you drink six or more standard alcoholic drinks on one occasion? | 1 Never (0 points) 2 Less than once a month (1 point) 3 Every month (2 points) 4 Every week (3 points) 5 Every day or almost every day (4 points) |  |  |  |
| **Health-related variables** | | | | | |
| BMI | Based on height and weight from clinical exam (kg/m^2^) |  | <18.5 18.5 to <25 25 to <30 30 or more | Most recent clinical exam | LCA |
| General health score | How do you rate your general state of health? | Very good: 1; 2; 3; 4; 5; 6; 7; 8: Very bad | Good: 1–2 Moderate: 3–4 Poor: ≥5 | 2020/2021 | LCA |
| Depression | Neurological and psychological disorders - Depression treated | 1 Yes 2 No |  | 2020/2021 | LCA |
| Abbreviation: body mass index (BMI), latent class analysis (LCA) | | | | | |
| Note: When variables were missing from the 2020/2021 questionnaire, the most recent value was used from questionnaires collected at follow-ups and baseline. | | | | | |
| *When answer categories are not specified, the same answer categories from the questionnaire were used. | | | | | |
| **Variables indicated as "LCA" were used to identify latent lifestyle profiles using LCA models. Variables indicated as "adjustment variable" were used as adjusment variables in logistic regression models. Some variables were used for both to reduce potential residual confounding. | | | | | |

**Supplemental Table 2. Demographic characteristics of women within the CRABAT cohort by latent lifestyle profile**

|  | **Profile 1** | | **Profile 2** | | **Profile 3** | | **Profile 4** | | **Profile 5** | | **Not assigned to a latent profile due to missing data*** | | **Overall** | |
| --- | --- | --- | --- | --- | --- | --- | --- | --- | --- | --- | --- | --- | --- | --- |
|  | (n=6,265) | | (n=5,270) | | (n=8,955) | | (n=10,073) | | (n=13,594) | | (n=6,612) | | (n=50,769) | |
|  | Older, not married | | Younger | | Older; married | | Risk-takers | | Socially-advantaged | |  |  |  |  |
|  | n | (%) | n | (%) | n | (%) | n | (%) | n | (%) | n | (%) | n | (%) |
| **Age** |  |  |  |  |  |  |  |  |  |  |  |  |  |  |
| <30 | 0 | (0%) | 1230 | (23%) | 0 | (0%) | 7 | (0%) | 0 | (0%) | 169 | (3%) | 1406 | (3%) |
| 30-39 | 50 | (1%) | 3114 | (59%) | 192 | (2%) | 2240 | (22%) | 2138 | (16%) | 926 | (14%) | 8660 | (17%) |
| 40-49 | 829 | (13%) | 853 | (16%) | 874 | (10%) | 4065 | (40%) | 5040 | (37%) | 1409 | (21%) | 13070 | (26%) |
| 50-59 | 2299 | (37%) | 73 | (1%) | 2722 | (30%) | 2422 | (24%) | 3952 | (29%) | 1704 | (26%) | 13172 | (26%) |
| 60-69 | 3087 | (49%) | 0 | (0%) | 5167 | (58%) | 1339 | (13%) | 2464 | (18%) | 2404 | (36%) | 14461 | (28%) |
| **Education** |  |  |  |  |  |  |  |  |  |  |  |  |  |  |
| No diploma | 1766 | (28%) | 241 | (5%) | 3583 | (40%) | 490 | (5%) | 0 | (0%) | 1420 | (21%) | 7500 | (15%) |
| High school diploma | 1313 | (21%) | 1467 | (28%) | 2782 | (31%) | 1053 | (10%) | 177 | (1%) | 892 | (13%) | 7684 | (15%) |
| College, Bachelor's diploma | 2642 | (42%) | 1976 | (37%) | 2584 | (29%) | 4861 | (48%) | 7267 | (53%) | 2209 | (33%) | 21539 | (42%) |
| Masters diploma or higher | 544 | (9%) | 1586 | (30%) | 6 | (0%) | 3669 | (36%) | 6150 | (45%) | 1273 | (19%) | 13228 | (26%) |
| Missing | 0 | (0%) | 0 | (0%) | 0 | (0%) | 0 | (0%) | 0 | (0%) | 818 | (12.4%) | 818 | (1.6%) |
| **Monthly income, in euros** |  |  |  |  |  |  |  |  |  |  |  |  |  |  |
| >1500 | 1449 | (23%) | 1405 | (27%) | 198 | (2%) | 339 | (3%) | 8 | (0%) | 705 | (11%) | 4104 | (8%) |
| >=1500-<2800 | 3495 | (56%) | 1992 | (38%) | 2270 | (25%) | 2158 | (21%) | 622 | (5%) | 1663 | (25%) | 12200 | (24%) |
| >=2800-<4200 | 957 | (15%) | 881 | (17%) | 4070 | (45%) | 3742 | (37%) | 4414 | (32%) | 1826 | (28%) | 15890 | (31%) |
| >=4200 | 97 | (2%) | 300 | (6%) | 1491 | (17%) | 3573 | (35%) | 8081 | (59%) | 1529 | (23%) | 15071 | (30%) |
| No answer | 267 | (4%) | 692 | (13%) | 926 | (10%) | 261 | (3%) | 469 | (3%) | 889 | (13%) | 3504 | (7%) |
| **Marital status** |  |  |  |  |  |  |  |  |  |  |  |  |  |  |
| Unmarried | 2526 | (40%) | 4875 | (93%) | 17 | (0%) | 2749 | (27%) | 1105 | (8%) | 1548 | (23%) | 12820 | (25%) |
| Married/civil partnership | 4 | (0%) | 390 | (7%) | 8825 | (99%) | 6417 | (64%) | 11736 | (86%) | 4122 | (62%) | 31494 | (62%) |
| Was married | 3735 | (60%) | 5 | (0%) | 113 | (1%) | 907 | (9%) | 753 | (6%) | 917 | (14%) | 6430 | (13%) |
| Missing | 0 | (0%) | 0 | (0%) | 0 | (0%) | 0 | (0%) | 0 | (0%) | 25 | (0.4%) | 25 | (0.0%) |
| **Ever smoking** |  |  |  |  |  |  |  |  |  |  |  |  |  |  |
| Yes | 3262 | (52%) | 1342 | (25%) | 4085 | (46%) | 9545 | (95%) | 2662 | (20%) | 2189 | (33%) | 23085 | (45%) |
| No | 3003 | (48%) | 3928 | (75%) | 4870 | (54%) | 528 | (5%) | 10932 | (80%) | 2932 | (44%) | 26193 | (52%) |
| Missing | 0 | (0%) | 0 | (0%) | 0 | (0%) | 0 | (0%) | 0 | (0%) | 1491 | (22.6%) | 1491 | (2.9%) |
| **Cannabis use** |  |  |  |  |  |  |  |  |  |  |  |  |  |  |
| Never | 5104 | (81%) | 2849 | (54%) | 8618 | (96%) | 176 | (2%) | 11796 | (87%) | 2972 | (45%) | 31515 | (62%) |
| Former | 1073 | (17%) | 1823 | (35%) | 327 | (4%) | 8888 | (88%) | 1783 | (13%) | 1371 | (21%) | 15265 | (30%) |
| Current | 88 | (1%) | 598 | (11%) | 10 | (0%) | 1009 | (10%) | 15 | (0%) | 173 | (3%) | 1893 | (4%) |
| Missing | 0 | (0%) | 0 | (0%) | 0 | (0%) | 0 | (0%) | 0 | (0%) | 2096 | (31.7%) | 2096 | (4.1%) |
| **Alcohol use** |  |  |  |  |  |  |  |  |  |  |  |  |  |  |
| Abstainer | 1820 | (29%) | 1577 | (30%) | 2312 | (26%) | 1085 | (11%) | 3017 | (22%) | 1382 | (21%) | 11193 | (22%) |
| Abuse or dependence | 467 | (7%) | 651 | (12%) | 581 | (6%) | 2224 | (22%) | 679 | (5%) | 579 | (9%) | 5181 | (10%) |
| Moderate use | 3978 | (63%) | 3042 | (58%) | 6062 | (68%) | 6764 | (67%) | 9898 | (73%) | 3735 | (56%) | 33479 | (66%) |
| Missing | 0 | (0%) | 0 | (0%) | 0 | (0%) | 0 | (0%) | 0 | (0%) | 916 | (13.9%) | 916 | (1.8%) |
| **BMI** |  |  |  |  |  |  |  |  |  |  |  |  |  |  |
| Underweight | 203 | (3%) | 443 | (8%) | 155 | (2%) | 477 | (5%) | 694 | (5%) | 243 | (4%) | 2215 | (4%) |
| Normal weight | 3359 | (54%) | 3507 | (67%) | 4290 | (48%) | 6836 | (68%) | 9786 | (72%) | 3548 | (54%) | 31326 | (62%) |
| Overweight | 1699 | (27%) | 856 | (16%) | 2869 | (32%) | 2008 | (20%) | 2451 | (18%) | 1497 | (23%) | 11380 | (22%) |
| Obese | 1004 | (16%) | 464 | (9%) | 1641 | (18%) | 752 | (7%) | 663 | (5%) | 829 | (13%) | 5353 | (11%) |
| Missing | 0 | (0%) | 0 | (0%) | 0 | (0%) | 0 | (0%) | 0 | (0%) | 495 | (7.5%) | 495 | (1.0%) |
| **Number of lifetime sexual partners** |  |  |  |  |  |  |  |  |  |  |  |  |  |  |
| None | 1718 | (27%) | 1777 | (34%) | 2544 | (28%) | 2718 | (27%) | 5699 | (42%) | 1829 | (28%) | 16285 | (32%) |
| One | 92 | (1%) | 578 | (11%) | 3 | (0%) | 0 | (0%) | 2 | (0%) | 119 | (2%) | 794 | (2%) |
| Two to five | 244 | (4%) | 741 | (14%) | 2727 | (30%) | 273 | (3%) | 3469 | (26%) | 980 | (15%) | 8434 | (17%) |
| Six or more | 1761 | (28%) | 1435 | (27%) | 546 | (6%) | 5245 | (52%) | 2099 | (15%) | 1177 | (18%) | 12263 | (24%) |
| No answer | 2450 | (39%) | 739 | (14%) | 3135 | (35%) | 1837 | (18%) | 2325 | (17%) | 2507 | (38%) | 12993 | (26%) |
| **Regular pap smears** |  |  |  |  |  |  |  |  |  |  |  |  |  |  |
| Yes | 4902 | (78%) | 3859 | (73%) | 7401 | (83%) | 8875 | (88%) | 12176 | (90%) | 5406 | (82%) | 42619 | (84%) |
| No | 1363 | (22%) | 1411 | (27%) | 1554 | (17%) | 1198 | (12%) | 1418 | (10%) | 1206 | (18%) | 8150 | (16%) |
| **At least one dose of HPV vaccine** |  |  |  |  |  |  |  |  |  |  |  |  |  |  |
| Yes | 49 | (1%) | 1585 | (30%) | 92 | (1%) | 405 | (4%) | 550 | (4%) | 333 | (5%) | 3014 | (6%) |
| No | 6216 | (99%) | 3685 | (70%) | 8863 | (99%) | 9668 | (96%) | 13044 | (96%) | 6279 | (95%) | 47755 | (94%) |

*Individuals were missing data for at least one LCA variable and therefore were unable to be categorized. These individuals were excluded in the regression analyses.

Abbreviations: Body mass index (BMI), human papillomavirus (HPV)

**Supplemental Table 3. Associations between latent lifestyle profile membership and ever receiving a tattoo among women under age 70**

|  |  | **Ever tattooed** (n=7,881) | | | **Never tattooed** (n=42,888) | | **Unadjusted** | **Adjusted** |
| --- | --- | --- | --- | --- | --- | --- | --- | --- |
|  |  | n | % | Tattoo prevalence | n | % | OR (95% CI) | OR (95% CI) |
| Latent lifestyle profile | |  |  |  |  |  |  |  |
| 1 | Older, not married | 915 | 12% | 15% | 5350 | 12% | 1.79 (1.63–1.96) | 1.65 (1.48–1.85) |
| 2 | Younger | 1439 | 18% | 27% | 3831 | 9% | 3.93 (3.61–4.28) | 0.94 (0.85–1.04) |
| 3 | Older, married | 923 | 12% | 10% | 8032 | 19% | 1.20 (1.10–1.32) | 1.01 (0.90–1.13) |
| 4 | Risk-takers | 2445 | 31% | 24% | 7628 | 18% | 3.35 (3.11–3.62) | 1.66 (1.51–1.82) |
| 5 | Socially-advantaged | 1186 | 15% | 9% | 12408 | 29% | Ref | Ref |

Models adjusted for age, ever smoking, and education level

**Supplemental Table 4. Associations between latent lifestyle profile membership and cervical cancer diagnosis among women under age 70**

|  | | **Excluding tattooed individuals** | | | | | **Including tattooed individuals** | | | | |
| --- | --- | --- | --- | --- | --- | --- | --- | --- | --- | --- | --- |
|  |  | **Cancer cases** (n=244) | | **Non-cases** (n=42,644) | | OR (95% CI) | **Cancer cases** (n=320) | | **Non-cases** (n=50,449) | | OR (95% CI) |
|  |  | n | % | n | % |  | n | % | n | % |  |
| Latent lifestyle profile | |  |  |  |  |  |  |  |  |  |  |
| 1 | Older, not married | 47 | 19% | 5,303 | 12% | 2.46 (1.58–3.87) | 58 | 18% | 6,207 | 12% | 2.47 (1.66–3.71) |
| 2 | Younger | 18 | 7% | 3,813 | 9% | 1.87 (0.99–3.42) | 24 | 8% | 5,246 | 10% | 1.31 (0.75–2.23) |
| 3 | Older, married | 37 | 15% | 7,995 | 19% | 1.32 (0.82–2.12) | 47 | 15% | 8,908 | 18% | 1.44 (0.94–2.20) |
| 4 | Risk-takers | 70 | 29% | 7,558 | 18% | 2.82 (1.91–4.20) | 99 | 31% | 9,974 | 20% | 2.61 (1.86–3.71) |
| 5 | Socially-advantaged | 40 | 16% | 12,368 | 29% | Ref | 50 | 16% | 13,544 | 27% | Ref |

Models adjusted for age, HPV vaccine, healthcare access, and regular Pap smears

**Supplemental Table 5. Tattooing exposures and risk of cervical cancer, limited to women receiving regular Pap smears**

|  | **Cancer cases** (n=269) | | **Non-cases** (n=42,350) | | Model 1 | Model 2 | Model 3 |
| --- | --- | --- | --- | --- | --- | --- | --- |
|  | n | % | n | % | OR (95% CI) | OR (95% CI) | OR (95% CI) |
| **Ever tattooed** |  |  |  |  |  |  |  |
| No | 204 | 76% | 35,683 | 84% | Ref | Ref | Ref |
| Yes | 65 | 24% | 6,667 | 16% | 1.43 (1.04–1.94) | 1.41 (1.02–1.93) | 1.38 (0.98–1.91) |
| **Tattooed body surface area** |  |  |  |  |  |  |  |
| Never tattooed | 204 | 76% | 35,683 | 84% | Ref | Ref | Ref |
| < One hand palm | 18 | 7% | 2,210 | 5% | 1.24 (0.72–2.00) | 1.20 (0.69–1.96) | 1.04 (0.56–1.77) |
| One hand palm or more | 24 | 9% | 1,934 | 5% | 1.94 (1.21–2.99) | 1.93 (1.20–2.99) | 1.89 (1.15–2.98) |
| **Tattoo context** |  |  |  |  |  |  |  |
| Never tattooed | 204 | 76% | 35,683 | 84% | Ref | Ref | Ref |
| Only tattooed by professional artist inside studio | 33 | 12% | 3,438 | 8% | 1.48 (0.99–2.16) | 1.46 (0.96–2.15) | 1.35 (0.86–2.03) |
| Tattooed in other circumstance | 8 | 3% | 666 | 2% | 1.81 (0.81–3.50) | 1.77 (0.79–3.45) | 1.64 (0.69–3.32) |
| **Time since first tattoo** |  |  |  |  |  |  |  |
| Never tattooed | 204 | 76% | 35,683 | 84% | Ref | Ref | Ref |
| Less than 10 years | 8 | 3% | 1,498 | 4% | 0.85 (0.38–1.63) | 0.79 (0.33–1.58) | 0.72 (0.28–1.51) |
| 10 or more years | 32 | 12% | 2,597 | 6% | 1.87 (1.24–2.74) | 1.84 (1.21–2.71) | 1.70 (1.09–2.57) |
| Missing data on timing of tattoo | 25 | 9% | 2,572 | 6% | - | - | - |

| Model 1 adjusted for age, ever smoking, and education |
| --- |
| Model 2 adjusted for age, ever smoking, education, number of lifetime sexual partners, HPV vaccination, and healthcare access |
| Model 3 adjusted for age, ever smoking, education, latent lifestyle profile, number of lifetime sexual partners, HPV vaccination, and healthcare access |

**Supplemental Table 6. Cox proportional hazards regression results for the association between tattoo exposure and cervical cancer risk, overall**

|  | **Model 1** | **Model 2** | **Model 3** |
| --- | --- | --- | --- |
|  | HR (95% CI) | HR (95% CI) | HR (95% CI) |
| **Approach 1: Ever/never** | | | |
| **Ever tattooed** |  |  |  |
| No | Ref | Ref | Ref |
| Yes | 1.48 (0.97–2.27) | 1.37 (0.90–2.10) | 1.20 (0.76–1.90) |
| **Approach 2: Time-varying** | | | |
| **Ever tattooed** |  |  |  |
| No | Ref | Ref | Ref |
| Yes | 1.53 (1.28–1.82) | 1.46 (1.22–1.75) | 1.35 (1.10–1.65) |
| **Tattoo exposure time** |  |  |  |
| Never tattooed | Ref | Ref | Ref |
| Tattooed in 1 time period | 3.24 (2.26–4.63) | 3.01 (2.10–4.30) | 2.72 (1.86–3.98) |
| Tattooed in 2+ time periods | 2.20 (1.11–4.37) | 2.02 (1.01–4.01) | 2.06 (1.03–4.10) |
| Tattooed in 3+ time periods | 1.66 (0.52–5.27) | 1.48 (0.47–4.69) | 0.51 (0.07–3.65) |

Model 1 adjusted for age, ever smoking, and education

Model 2 adjusted for age, ever smoking, education, number of lifetime sexual partners, HPV vaccination, healthcare access, and regular Pap smears

Model 3 adjusted for age, ever smoking, education, latent lifestyle profile, number of lifetime sexual partners, HPV vaccination, healthcare access, and regular Pap smears

Note: Approach 2 uses the five retrospective tattoo timing categories as time-varying exposure updates. Each reported timing category contributed a split point in the follow-up record at its estimated midpoint date, incrementing a cumulative count of reported tattoo periods. This variable was categorized as never tattooed (0 periods), tattooed in one reported period, two reported periods, or three or more reported periods for presentation.

**Supplemental Table 7. Cox proportional hazards regression results for the association between tattoo exposure and cervical cancer risk, stratified by latent lifestyle profile**

|  | **Class 1**  Older, not married | **Class 2**  Younger | **Class 3**  Older, married | **Class 4**  Risk-takers | **Class 5**  Socially-advantaged |
| --- | --- | --- | --- | --- | --- |
| **Approach 1: Ever/never** | | | | | |
| **Ever tattooed** |  |  |  |  |  |
| No | Ref | Ref | Ref | Ref | Ref |
| Yes | 1.23 (0.47–3.25) | 1.16 (0.39–3.43) | 1.65 (0.47–5.75) | 1.18 (0.54–2.56) | 1.56 (0.47–5.24) |
| **Approach 2: Time-varying** | | | | | |
| **Ever tattooed** |  |  |  |  |  |
| No | Ref | Ref | Ref | Ref | Ref |
| Yes | 1.33 (0.85–2.08) | 1.10 (0.63–1.93) | 1.25 (0.58–2.69) | 1.49 (1.11–2.01) | 1.80 (1.11–2.93) |

**Supplemental Table 8. E-values calculated for ORs from Model 3 (corresponding with Table 3)**

|  | Model 3 | E-value |
| --- | --- | --- |
|  | **OR (95% CI)** |  |
| **Ever tattooed** |  |  |
| No | Ref |  |
| Yes | 1.29 (0.95–1.73) | 1.90 |
| **Tattooed body surface area** |  |  |
| Never tattooed | Ref |  |
| < One hand palm | 1.03 (0.60–1.65) | 1.21 |
| One hand palm or more | 1.58 (0.99–2.43) | 2.54 |
| **Tattoo context** |  |  |
| Never tattooed | Ref |  |
| Only tattooed by professional artist inside studio | 1.28 (0.85–1.61) | 1.88 |
| Tattooed in other circumstance | 1.24 (0.52–2.49) | 1.79 |
| **Time since first tattoo** |  |  |
| Never tattooed | Ref |  |
| Less than 10 years | 0.86 (0.40–1.60) | 1.59 |
| 10 or more years | 1.43 (0.94–2.11) | 2.21 |
| Missing data on timing of tattoo | - |  |

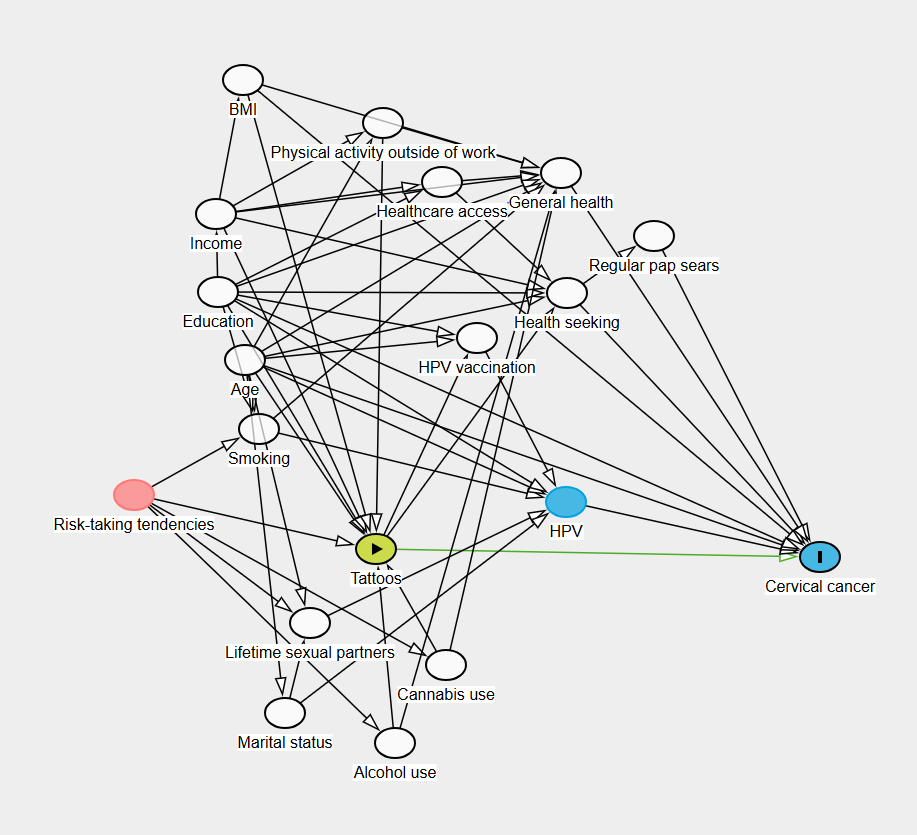

**Legend**

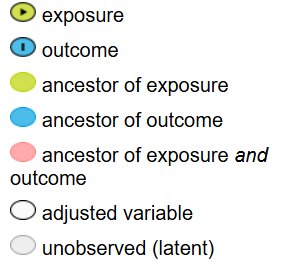

**Supplemental Figure 1.** The directed acyclic graph (DAG) used to visualize associations between tattoos and cervical cancer diagnosis and potential confounders.

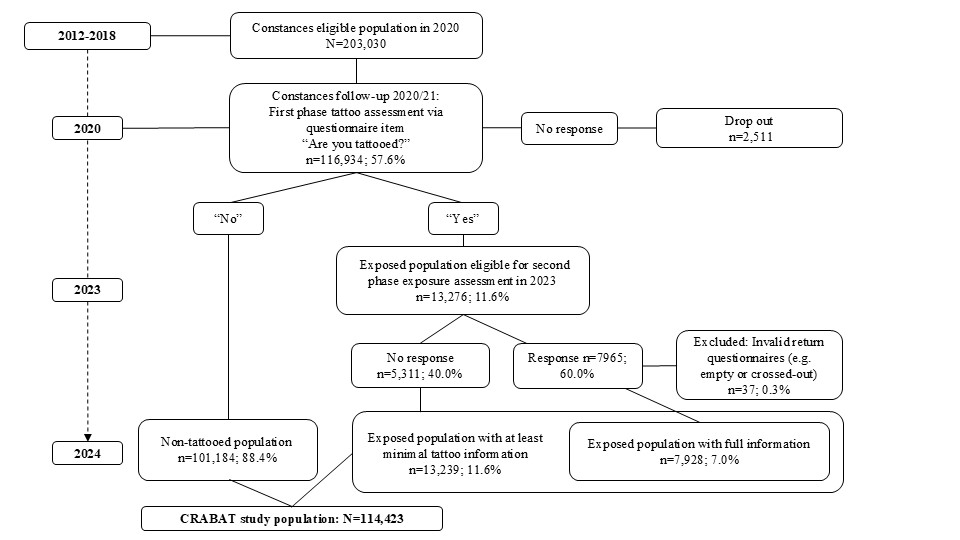

Supplemental Figure 2. Flow chart of sample determination of the CRABAT cohort.

Adapted from Hosseini B, McCarty R, Zins M, et al. Cohort Profile: The Cancer Risk Attributable to the Body Art of Tattooing (CRABAT) study. Int J Epidemiol. 2025;54(4):dyaf132. doi:10.1093/ije/dyaf132.
